# Spatial and Machine Learning Analysis of Breast and Cervical Cancer Screening Uptake in Ghana: Evidence from the 2022 Ghana Demographic and Health Survey

**DOI:** 10.64898/2026.08.15.26360510

**Authors:** Abubakar Hudu Siddick, Abubakar Iddrisu Siddiq, Osman Abdul-Fatawu Iddrisu

**Affiliations:** Department of Statistics and Actuarial Science, University of Ghana, Accra, Ghana; Department of Public Health, Faculty of Medical Sciences, Presbyterian University, Agogo, Ghana; Department of Epidemiology and Biostatistics, Kwame Nkrumah University of Science and Technology, Kumasi, Ghana

**Keywords:** Breast cancer screening, cervical cancer screening, Ghana, Demographic and Health Survey, spatial autocorrelation, machine learning, concentration index

## Abstract

**Background:** Breast and cervical cancer screening in Ghana remains low, and several analyses of the Ghana Demographic and Health Survey (GDHS) have already shown that wealth, education, and place of residence pattern who gets screened [1–3]. Whether this patterning clusters geographically below the level of administrative region has not been tested for this population, and whether cluster-aware machine learning adds anything to the standard regression approach used so far remains open.

**Methods:** We analyzed the 2022 Ghana Demographic and Health Survey women’s file (N = 15,014; primary sample of women aged 25-49 years, n = 9,510) linked to cluster geographic coordinates for 618 enumeration areas. Clinical breast examination and cervical cancer testing were the two outcomes. We estimated survey-weighted prevalence across demographic and socioeconomic strata, tested global spatial autocorrelation with Moran’s I, mapped local clustering with Getis-Ord Gi* statistics, and separately fitted gradient-boosted classifiers on individual-level socioeconomic covariates, validated under cluster-held-out five-fold cross-validation to prevent within-cluster information leakage. Feature contributions to the breast-screening model were interpreted with an additive, feature-level explanation technique, and socioeconomic inequality was quantified with both the ordinary and Erreygers-corrected concentration index. The predictive models did not include geographic coordinates or survey weights; both are noted as limitations.

**Results:** Weighted prevalence among women aged 25-49 years was 22.5% (standard error 0.77) for breast examination and 6.9% (standard error 0.46) for cervical testing. Both rose with education and wealth and were roughly double in urban areas relative to rural ones. Moran’s I was positive and significant for both outcomes (breast: 0.217, z = 11.69, p < 0.001; cervical: 0.125, z = 6.77, p < 0.001), and local cluster statistics located discrete hotspots around Greater Accra and parts of Ashanti and Bono, with coldspots concentrated across the north, though these local tests were not adjusted for multiple comparisons. Cross-validated discrimination reached an area under the curve of 0.716 for breast examination and 0.730 for cervical testing, without confidence intervals or calibration assessment; education, wealth, and age were the dominant predictors for both. The Erreygers index put breast examination as the more wealth-concentrated outcome (0.231 versus 0.095 for cervical testing), reversing the ranking implied by the uncorrected index.

**Conclusions:** Screening uptake in Ghana is spatially clustered at a resolution that regional reporting cannot show, and this clustering is compositionally associated with, though not formally shown to be mediated by, the socioeconomic makeup of individual clusters. Cluster-level spatial analysis and a model-based risk ranking may offer a useful complement to regional targeting, but calibration, external geographic validation, and comparison against a regional-allocation baseline are needed before any operational use.

**Trial registration:** Not applicable. This is a secondary, hypothesis-generating cross-sectional analysis of existing, publicly available survey data and was not prospectively registered.

## Background

Breast and cervical cancer together account for a large and rising share of the female cancer burden in sub-Saharan Africa [4,5]. In settings without population-wide mammography or HPV-based testing, clinical breast examination and opportunistic cervical screening remain the main tools available for early detection, and the World Health Organization has identified scaling up screening coverage as central to its global strategy for cervical cancer elimination [6]. In Ghana, uptake of both services is low. The 2022 round of the Ghana Demographic and Health Survey (GDHS) has already been analyzed several times for this question, and a consistent picture has emerged: wealth, education, health insurance status, and age predict screening uptake, and urban women in wealthier households are screened far more often than rural or poorer women [1–3,7]. Regional variation has also been documented, along with growing interest in the role that distance to a health facility plays as a structural barrier, particularly for cervical screening [8–10].

What none of this work has done is treat screening as a spatial phenomenon below the level of the survey’s administrative regions. The GDHS reports at most fourteen to sixteen regions, yet the sampling frame provides geographic coordinates for all 618 primary sampling clusters, albeit randomly displaced for confidentiality [11], and screening access plausibly follows facility catchment and local health-system capacity in ways that do not respect regional lines. A cluster in northern Ashanti may resemble a cluster in neighboring Bono more than it resembles a cluster near Kumasi. Small-area estimation that stops at the regional level risks averaging away exactly the local heterogeneity that a targeting exercise would need.

A second gap is methodological. Ghanaian studies of breast and cervical screening using the GDHS have relied on standard or multilevel logistic regression [2,12,13], which imposes a linear, additive functional form. Gradient-boosted decision trees can capture non-linear and interaction effects among covariates without a pre-specified functional form [14], and this approach has already been used to predict cervical and breast screening uptake elsewhere in sub-Saharan Africa and in other low-and middle-income settings [15–19]. A separate concern is validation: ordinary cross-validation on clustered survey data can overstate how well a model generalizes, because women sampled from the same cluster share unobserved local conditions (the same clinic, the same outreach history) that leak information between training and test folds when clusters, not individuals, are the true sampling unit [20]. Assigning whole clusters, rather than individual women, to cross-validation folds removes this specific leakage. It is important to be precise about what this design does and does not establish: it tests whether a model generalizes to women in clusters not used for training, given that the model’s own covariates contain no geographic information; it does not test extrapolation to geographically distant or contiguous unsampled regions, since neighboring clusters can still be split across folds by chance.

A small number of recent studies have applied spatial and machine-learning methods to related outcomes in Ghana, including health insurance coverage [21] and antenatal care access [22], and to cancer-screening spatial patterns elsewhere on the continent ([23], for Tanzania; [24], for Ghanaian young women’s health screening broadly). Cancer screening specifically has not, to our knowledge, been examined this way for Ghana.

This study combines two analytic strands that are complementary but distinct, and we keep them explicitly separate throughout. The first is a genuine spatial analysis: Moran’s I and Getis-Ord Gi* statistics test whether cluster-level screening rates are spatially autocorrelated and, if so, where statistically significant hotspots and coldspots fall. The second is an individual-level predictive analysis: cluster-held-out, cross-validated gradient-boosted models estimate each woman’s probability of screening from socioeconomic and demographic covariates, none of which include geographic coordinates. The two analyses are linked only through the cluster-level aggregation used to build a risk-ranking of clusters, and the manuscript should not be read as claiming an integrated spatial-machine-learning model. Breast examination and cervical testing are analyzed with the same sample, covariates, and validation design throughout, so that any difference between them reflects the outcomes rather than inconsistencies in method. Inequality in both outcomes is additionally quantified with the concentration index, reporting both the ordinary and the Erreygers-corrected version, since the ordinary index for a binary outcome is bounded by the outcome’s own mean prevalence and can therefore misstate the comparison between two outcomes with very different baseline rates [25].

We position this study as a methodological extension of an already-documented finding, not a rediscovery of it: Ghanaian women’s screening uptake is patterned by wealth and education. Our question is whether adding cluster-resolution spatial description and a validated predictive layer to that finding changes what a program planner could do with it, while being explicit about what remains untested among other things, calibration, geographic transportability beyond the sampled clusters, and performance relative to conventional regression.

## Methods

This is a secondary, hypothesis-generating cross-sectional analysis and was not prospectively registered. This study is not a randomized controlled trial, so CONSORT 2010 reporting guidance does not apply. Reporting instead follows the STROBE guideline for cross-sectional studies [26] where applicable to a secondary survey analysis; items concerning participant flow, missing data, and statistical uncertainty that are not fully resolved here are flagged explicitly in the in section 5.

### Data source and analytic sample

Data came from the individual (women’s) recode of the 2022 GDHS, a nationally representative two-stage cluster household survey, together with the companion geographic dataset giving displaced Global Positioning System (GPS) coordinates for each of the survey’s 618 clusters [11], linked by cluster identifier. The full recode contained 15,014 women with non-missing responses on both screening outcomes. The primary analytic sample was restricted to women aged 25 to 49 years (n = 9,510), matching the age band used in comparable Ghanaian analyses of this survey round [13,27]. This restriction was adopted for comparability with prior work rather than derived from a formal power calculation, and we did not conduct a separate a priori sample-size or power analysis for the spatial tests, subgroup estimates, or machine-learning models.

### Outcomes and covariates

The two outcomes, clinical breast examination and cervical cancer testing, were binary indicators of ever having received each service, taken directly from the corresponding GDHS items; both are self-reported, ever-use indicators rather than verified or recent screening. Covariates were age in completed years, household wealth quintile, educational attainment (none, primary, secondary, higher), urban or rural residence, health insurance coverage, two barrier indicators (distance to a facility and money needed for treatment, each reported as a big problem), a media-exposure indicator (regular newspaper, radio, or television exposure), and region of residence entered as a set of indicator variables. Records with missing values on any covariate were dropped case-wise from the machine-learning sample; the number and pattern of records excluded this way, and how the excluded women compared with those retained, were not formally tabulated in this analysis.

### Survey-weighted descriptive statistics

All prevalence estimates use the GDHS sampling weights and account for stratification and clustering. A survey design object was built from individual sample weights, cluster (primary sampling unit), and strata, and weighted means with linearized standard errors were computed for both outcomes overall and within age group, education, wealth quintile, residence, and region. Cluster-level rates used for the spatial analysis below, by contrast, were computed as unweighted within-cluster means of the individual outcome; we did not carry the individual sampling weights into the cluster aggregation, so the reported cluster rates and the Moran’s I and Gi* statistics built from them should be read as descriptive of the sampled women in each cluster rather than as formally survey-weighted small-area estimates.

### Spatial autocorrelation and hotspot detection

Cluster-level screening rates were built by aggregating individual outcomes within each of the 618 clusters, keeping clusters with at least five sampled women aged 25-49; this threshold followed the convention used in comparable DHS-based spatial screening analyses [23] rather than a formal optimization, and we did not test alternative thresholds. A spatial weights matrix was built from k-nearest-neighbor adjacency (k = 8) with row-standardized weights wij; k = 8 was fixed a priori rather than selected by sensitivity analysis, and alternative neighborhood definitions (different k, distance-based weights) could alter the precise clusters flagged as significant, which we note as a limitation. Global spatial autocorrelation in the cluster rate xi was tested with Moran’s I [28]:

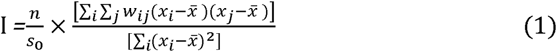

where n is the number of clusters, x□ is the mean cluster rate, and S_0_=Σ*_i_*Σ*_j_w_i,j_* is the sum of all spatial weights. Under the randomization assumption, I has an expected value of −1/(n−1) and a variance available in closed form [28]; departures from this expectation are converted to a z-score for a two-sided test of non-random spatial pattern. We used the conventional randomization-based inference implemented in the spdep package and did not additionally verify this assumption against a permutation-based null distribution using the unweighted, unequal-sample-size cluster rates described above.

Local clustering was mapped with the Getis-Ord Gi* statistic [29], computed at each cluster i over the same neighbor set:

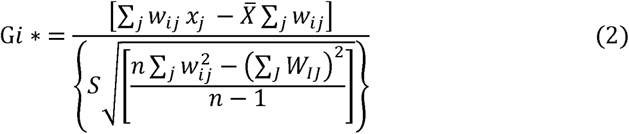

where X□ and S are the mean and standard deviation of *x_i_* over all clusters. Clusters with Gi* > 1.96 were classified as statistically significant hotspots and clusters with Gi* <-1.96 as coldspots, at approximately the 5% level per test. Because Gi* was evaluated at every eligible cluster simultaneously (several hundred local tests per outcome), the number of clusters flagged as significant by chance alone is expected to exceed 5% of the true null cases; we did not apply a false-discovery-rate or other multiplicity correction, so the hotspot and coldspot locations reported in this paper should be read as exploratory rather than confirmatory, and we flag this explicitly rather than implying formal inferential control.

### Individual-level predictive modeling and cluster-held-out validation

Gradient-boosted classifiers were fitted separately for each outcome using age, wealth, education, urban residence, insurance status, the two barrier indicators, media exposure, and region dummies as features. Geographic coordinates were not included as predictors, so these models are individual-level socioeconomic prediction models rather than spatial prediction models; we use that description throughout rather than referring to them as spatial machine learning. Extreme Gradient Boosting (XGBoost) builds an additive ensemble of regression trees fk by minimizing a regularized objective [14]:

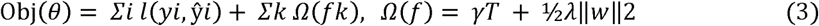

where l is the logistic loss between the observed outcome *y_i_* and the predicted probability,*ŷ_i_* T is the number of leaves in a tree, w the leaf weights, and γ and λ the complexity and L2 regularization penalties. Individual sampling weights were not incorporated into model fitting (XGBoost supports per-observation weights, but this option was not used here); consequently, the reported area under the curve (AUC) and feature-importance values describe discrimination and predictor contribution within the achieved analytic sample and should not be interpreted as formally survey-weighted estimates for the national target population.

Because women from the same cluster are not independent draws, ordinary k-fold cross-validation, which can place women from one cluster in both the training and the test fold, would overstate generalization to unseen women. Five-fold cross-validation was instead constructed so that clusters, not individual women, were the unit randomly assigned to folds (via the caret::createFolds on the vector of unique cluster identifiers), guaranteeing that every woman from a given cluster fell entirely in either the training or the test partition within a fold. This prevents within-cluster leakage; it does not, by itself, place geographically separated or contiguous clusters into different folds, since fold assignment was a random partition of clusters rather than a geographically stratified or spatially contiguous blocking scheme. We therefore refer to this design as cluster-held-out cross-validation rather than geographically blocked cross-validation, and we did not additionally test a leave-region-out or spatially contiguous blocking scheme, which would give a stricter test of geographic extrapolation. Each fold trained a binary-logistic XGBoost model (maximum depth 4, learning rate 0.05, 80% row and column subsampling, minimum child weight 5, with early stopping on the held-out fold’s AUC after 30 non-improving rounds) on the remaining four folds and evaluated it on the held-out clusters; because the held-out fold served as both the early-stopping criterion and the performance-evaluation set within each fold. The reported fold AUCs may carry a small optimistic bias from this shared use, which an inner validation split would avoid. We report the mean AUC across the five folds as the primary metric, together with the AUC from pooling all out-of-fold predictions into a single receiver operating characteristic (ROC) curve. Neither is accompanied by a confidence or bootstrap interval, and we did not compute one, which limits comparison between the two outcomes’ point estimates. Final models used for importance and SHAP analysis were refit on the complete analytic sample with the same hyperparameters over 250 boosting rounds rather than the per-fold early stopping rule, which introduces a further source of ambiguity about the exact number of boosting rounds represented in the importance and SHAP results. Hyperparameters (tree depth, learning rate, subsampling fractions, minimum child weight) were fixed a priori rather than tuned by nested cross-validation, and no comparison against a survey-weighted logistic-regression or multilevel-regression baseline was performed. A global random seed (20260808) was set at the start of the analysis, governing fold assignment among other steps, and a separate seed (42) was set before the SHAP subsampling step; package and R version numbers were not recorded in the analysis log.

### Feature importance, SHAP, and cluster risk ranking

Feature importance was summarized with XGBoost’s gain metric, the average loss reduction attributable to splits on a given feature across all trees. For the breast-examination model, Shapley additive explanations (SHAP) values [30] were computed on a random subsample of 2,000 women. Each woman’s prediction is decomposed into additive, feature-level contributions that show both the direction and the size of each covariate’s effect. These are predictive associations, not causal or mediating effects, and we use that language throughout rather than describing any covariate as mediating the outcome. Predicted probabilities from the final models were then averaged to the cluster level and used to rank clusters by mean predicted non-uptake, producing a composition-based ranking of the sampled clusters. Because this ranking is built from individual-level socioeconomic predictions rather than from a model with cluster-level spatial predictors, it reflects each cluster’s socioeconomic composition rather than an independent geographic prediction. We did not quantify uncertainty in the ranking (for example, via bootstrap resampling of clusters) and did not compare it against a regional-allocation baseline for targeting accuracy, calibration, or decision utility, all of which we treat as necessary before any programmatic use.

### Concentration index

Wealth-related inequality in each outcome was quantified with the concentration index [31]. Women were ranked by household wealth using a weight-consistent fractional rank *r_i_*, assigning each woman the midpoint of the cumulative weighted population share she occupies; ties in wealth rank were broken by the order in which records appeared in the sorted data, and strata and clusters were not separately incorporated into the covariance estimator beyond their role in the sampling weights themselves. The concentration index is twice the weighted covariance between the outcome yi and this rank, normalized by the weighted mean μ:

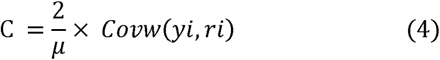

Because the ordinary index for a 0/1 outcome is bounded by 1 − μ, it mechanically compresses toward zero for a rare outcome such as cervical testing (μ = 0.069) relative to a more common one such as breast examination (μ = 0.225), making the two indices non-comparable on their raw scale. The Erreygers correction [25] improves comparability across binary outcomes with different prevalence by rescaling:

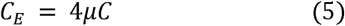

which places both outcomes’ indices on the same −1 to +1 scale. This correction depends on the specific normalization Erreygers proposed and on the wealth-ranking and weighting choices described above. Other inequality indices (for example, the Wagstaff normalization or the slope index of inequality) could yield a different comparison, and we did not cross-check the result against an alternative index. Neither index was accompanied by a standard error or bootstrap confidence interval, so the direction of the reversal between the ordinary and Erreygers-corrected comparison should be read as a point estimate rather than a statistically confirmed difference. Concentration curves, plotting the cumulative share of screening against the cumulative wealth-ranked population share, were plotted alongside both indices; a curve below the diagonal of equality indicates pro-rich concentration.

### Software and reproducibility

All analyses were run in R. Survey-weighted estimation used the survey package; spatial weights, Moran’s I, and Getis-Ord Gi* used spdep; gradient boosting used xgboost with cross-validation folds built in caret; SHAP values used shapviz; and administrative boundaries for mapping were obtained from GADM. Exact package and R versions were not captured in the original analysis log; a revised submission will include full sessionInfo() output. Analysis code is available from the corresponding author on request.

### Use of generative AI tools

Generative artificial intelligence (AI) and large language model tools were used to assist with formatting this manuscript to the target journal’s structural and reference-style requirements and with language editing during manuscript preparation. No generative AI tool was used to design the study, collect or analyze data, generate results, or draw scientific conclusions, and no such tool is listed as an author or is credited with responsibility for the scientific content of this work; the authors take full responsibility for the accuracy and integrity of the manuscript.

## Results

### Sample and overall prevalence

The primary sample comprised 9,510 women aged 25-49 with non-missing outcome data. The further reduction to the complete-case machine-learning sample after dropping records with any missing covariate is not separately reported here. Weighted prevalence of clinical breast examination was 22.5% (standard error 0.77) and of cervical cancer testing was 6.9% (standard error 0.46), broadly consistent with, though somewhat higher than, national estimates reported for the wider 15-49 age range in the same survey round [1]. Regional sample sizes ranged from roughly 490 to 770 women per region, even enough that the regional prevalence differences reported below are not simply an artifact of sample-size imbalance.

### Socioeconomic and demographic patterning

Both outcomes rose across every socioeconomic gradient examined (Figure 1). By education, breast-examination prevalence rose from 9.9% among women with no formal schooling to 48.7% among women with higher education; cervical testing rose from 3.2% to 19.7% over the same categories (Table 1). By wealth quintile, breast examination rose from 9.8% (poorest) to 39.5% (richest), and cervical testing from 2.0% to 13.0%. These exact, survey-weighted figures are given in Table 1.

**Figure 1.**
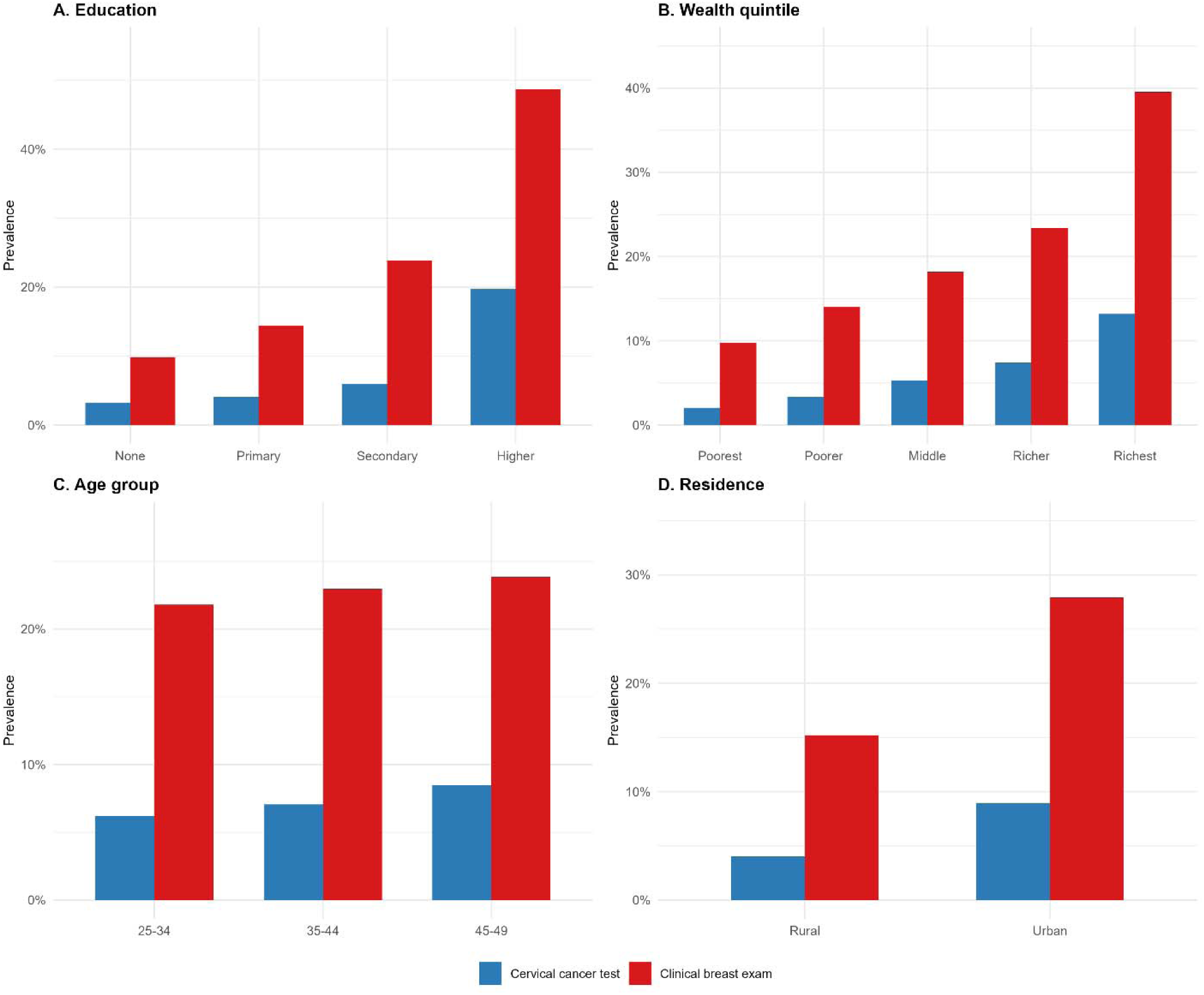
Weighted screening prevalence by education, wealth quintile, age group, and residence, women 25-49 years, Ghana DHS 2022.

**Table 1.** Weighted screening prevalence by education and wealth quintile, women 25-49 years, Ghana Demographic and Health Survey 2022.

| Stratum | Breast exam (%) | Cervical test (%) |
| --- | --- | --- |
| Education: None | 9.9 | 3.2 |
| Education: Primary | 14.4 | 4.1 |
| Education: Secondary | 23.9 | 6.0 |
| Education: Higher | 48.7 | 19.7 |
| Wealth: Poorest | 9.8 | 2.0 |
| Wealth: Poorer | 14.0 | 3.4 |
| Wealth: Middle | 18.2 | 5.3 |
| Wealth: Richer | 23.4 | 7.4 |
| Wealth: Richest | 39.5 | 13.0 |

Regional prevalence of clinical breast examination ranged from 8.6% to 29.6%, more than a three-fold gap that a regional average alone does not resolve geographically (Figure 2); the sections below examine this variation at cluster resolution.

**Figure 2.**
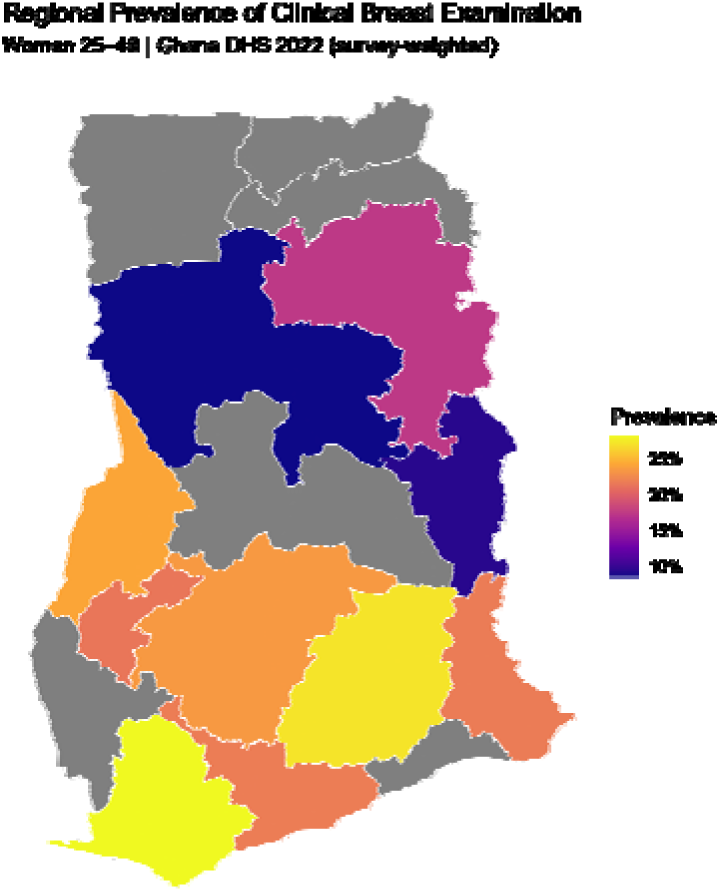
Regional prevalence of clinical breast examination, women 25-49 years. Grey regions had insufficient boundary-matched cluster data for shading.

### Spatial autocorrelation and hotspots

Global Moran’s I, computed on unweighted cluster-level rates, indicated significant positive spatial autocorrelation for both outcomes: breast examination I = 0.217 (z = 11.69, p < 0.001), cervical testing I = 0.125 (z = 6.77, p < 0.001). The larger coefficient for breast examination suggests a stronger, more geographically contiguous pattern than for cervical testing, though both departures from spatial randomness were significant under the randomization test used. We did not additionally check this result against a permutation null built from survey-weighted or stabilized cluster rates, so these coefficients should be read as descriptive of the raw cluster sample rather than as a fully design-consistent small-area estimate.

Getis-Ord Gi* statistics localized this clustering to specific clusters (Figures 3-4). For breast examination, apparent hotspots concentrated around Greater Accra, an Ashanti-Bono border area, and pockets in the Western and Central regions, while apparent coldspots concentrated across the Northern, North East, Upper East, Upper West, and parts of the Eastern region.

**Figure 3.**
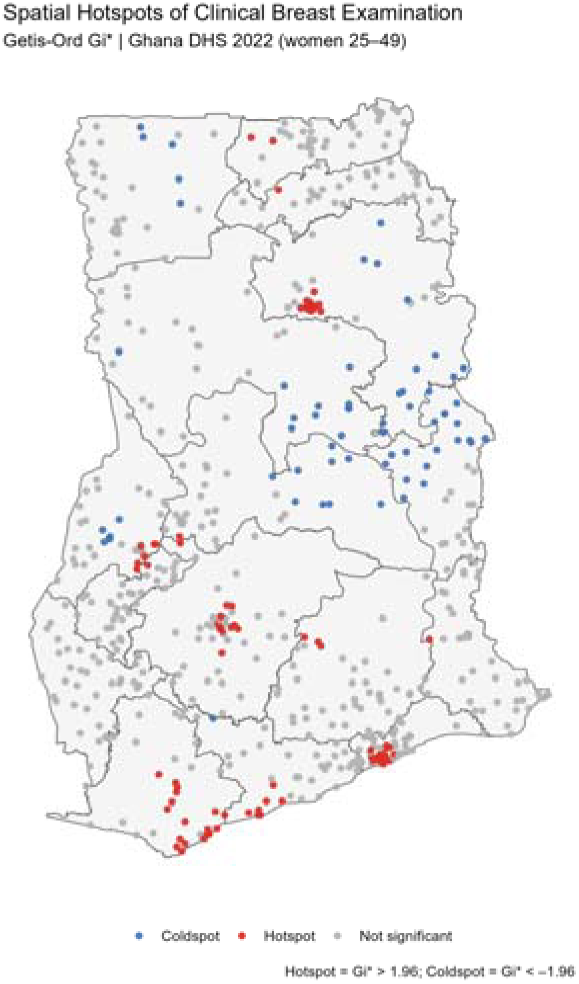
Getis-Ord Gi* local statistics for clinical breast examination uptake, by cluster (exploratory; not adjusted for multiple comparisons).

**Figure 4.**
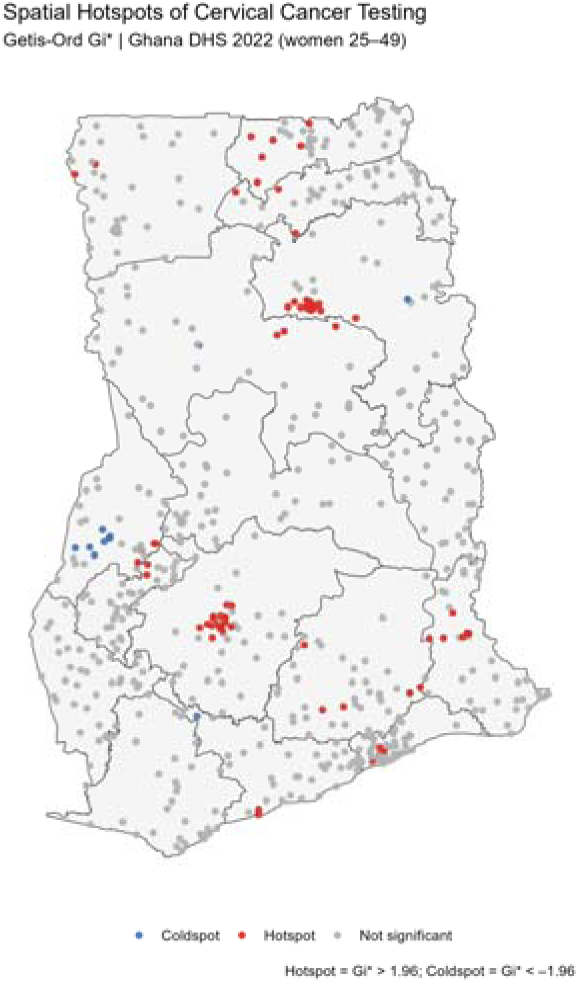
Getis-Ord Gi* local statistics for cervical cancer testing uptake, by cluster (exploratory; not adjusted for multiple comparisons).

Cervical-testing hotspots followed a broadly similar southern concentration but extended further north than breast-examination hotspots did, suggesting the two services’ spatial reach is correlated but not identical, consistent with facility-distance patterns reported for cervical screening elsewhere in the subregion [9]. Because several hundred local Gi* tests were run per outcome without a multiple-comparison correction, the precise number and boundary of clusters we describe as significant should be treated as an exploratory map rather than a confirmed set of hotspots. We did not tabulate the exact count of significant clusters for this reason and flag formal multiplicity control as a priority for a further analysis.

### Predictive performance

Under cluster-held-out five-fold cross-validation, mean AUC was 0.716 for breast examination and 0.730 for cervical testing (Table 2). Pooling out-of-fold predictions into a single ROC curve gave 0.704 and 0.716 respectively (Figure 5). Both figures indicate moderate discrimination for both outcomes; we do not interpret the closeness of the fold-averaged and pooled estimates as evidence against overfitting. Because the held-out fold in each iteration was used both for early stopping and for performance evaluation, some optimism cannot be ruled out, and no confidence interval accompanies either AUC. Comparable machine-learning models of cervical screening elsewhere in the region have reported AUCs in a broadly similar range [15,16], though direct comparison is limited by differences in covariates, validation design, and outcome definition across studies.

**Figure 5.**
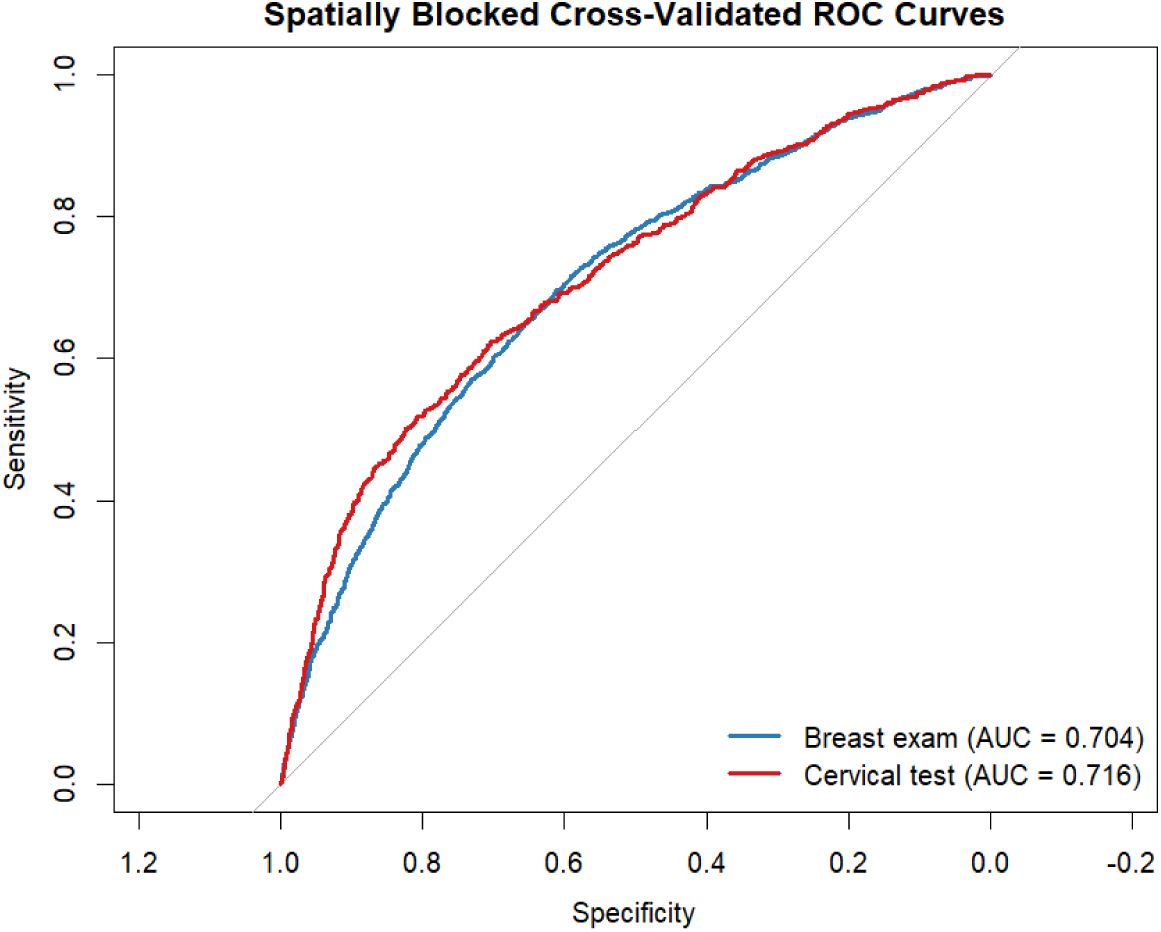
Cluster-held-out, cross-validated ROC curves for clinical breast examination and cervical cancer testing.

**Table 2.** Predictive performance (discrimination only, no confidence intervals or calibration) and global spatial autocorrelation, by outcome.

| Outcome | Mean fold AUC (5-fold cluster-held-out CV) | Pooled out-of-fold ROC AUC | Global Moran's I (cluster rate) |
| --- | --- | --- | --- |
| Clinical breast examination | 0.716 | 0.704 | 0.217 ($p < 0.001$) |
| Cervical cancer testing | 0.730 | 0.716 | 0.125 ($p < 0.001$) |

### Feature importance and SHAP

Feature-importance rankings (Figure 6) were consistent across outcomes: education was the top predictor for both (gain approximately 0.36 for breast examination, 0.31 for cervical testing), followed by wealth and age, which traded second and third rank between outcomes. Media exposure and reported money problems contributed modestly, and individual region indicators contributed comparatively little relative to the socioeconomic covariates. Because the models contain no geographic coordinates, this last observation means only that, among the covariates supplied, region membership itself adds little beyond individual socioeconomic composition; it does not by itself explain the spatial clustering documented above, which would require a formal decomposition or mediation analysis that we did not conduct.

**Figure 6.**
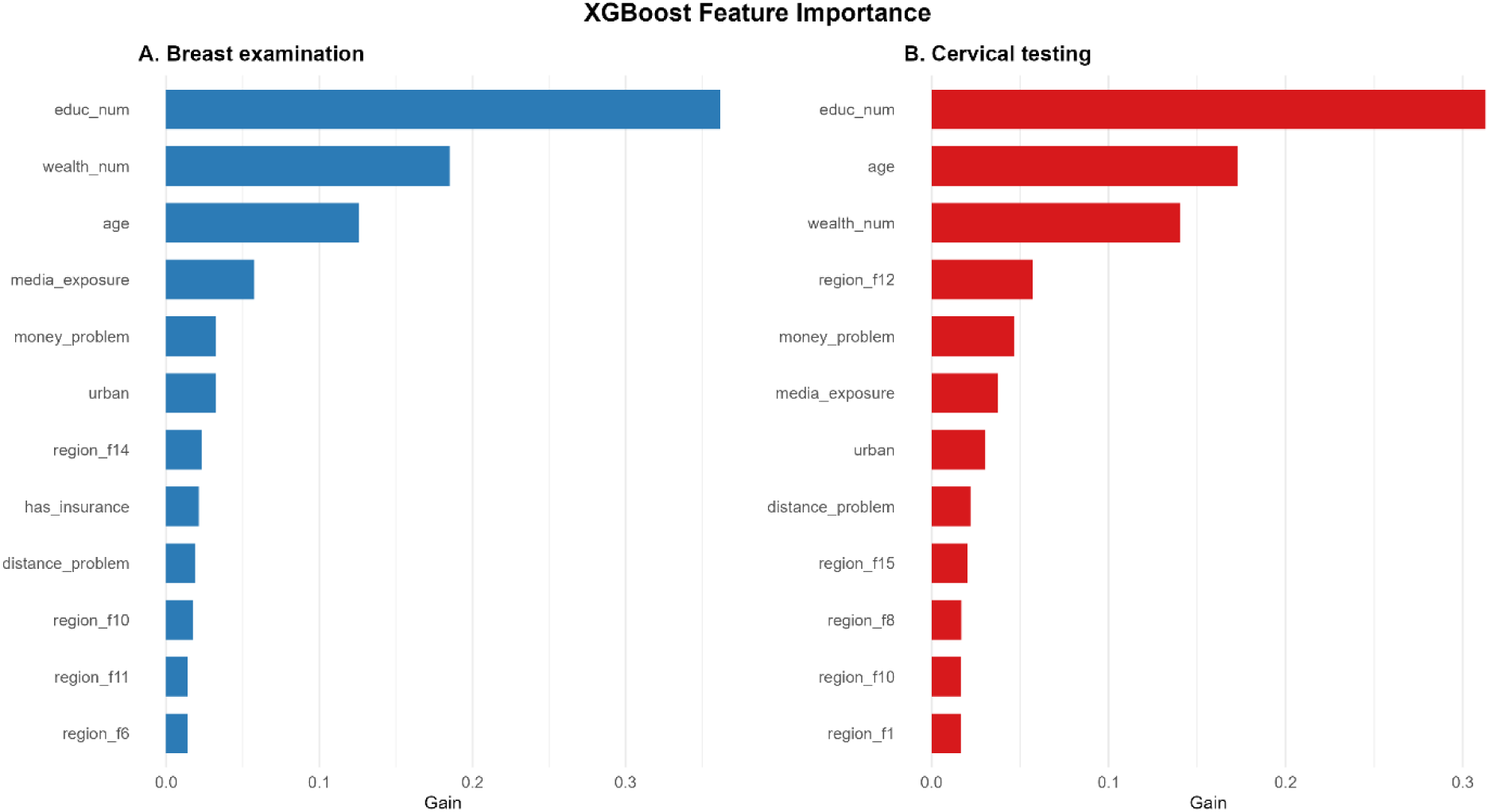
XGBoost feature importance (gain) for clinical breast examination and cervical cancer testing, cluster-held-out models.

The SHAP summary for breast examination (Figure 7) shows high education and high wealth pushing predicted probability up sharply and across a wide range, while lacking insurance had a smaller but consistently negative contribution. These are predictive associations within the fitted model and should not be read as causal or mediating effects. A subset of region indicators showed a right-skewed spread of positive SHAP values for a minority of women, a pattern that is consistent with, but does not itself confirm, the discrete localized hotspots identified above.

**Figure 7.**
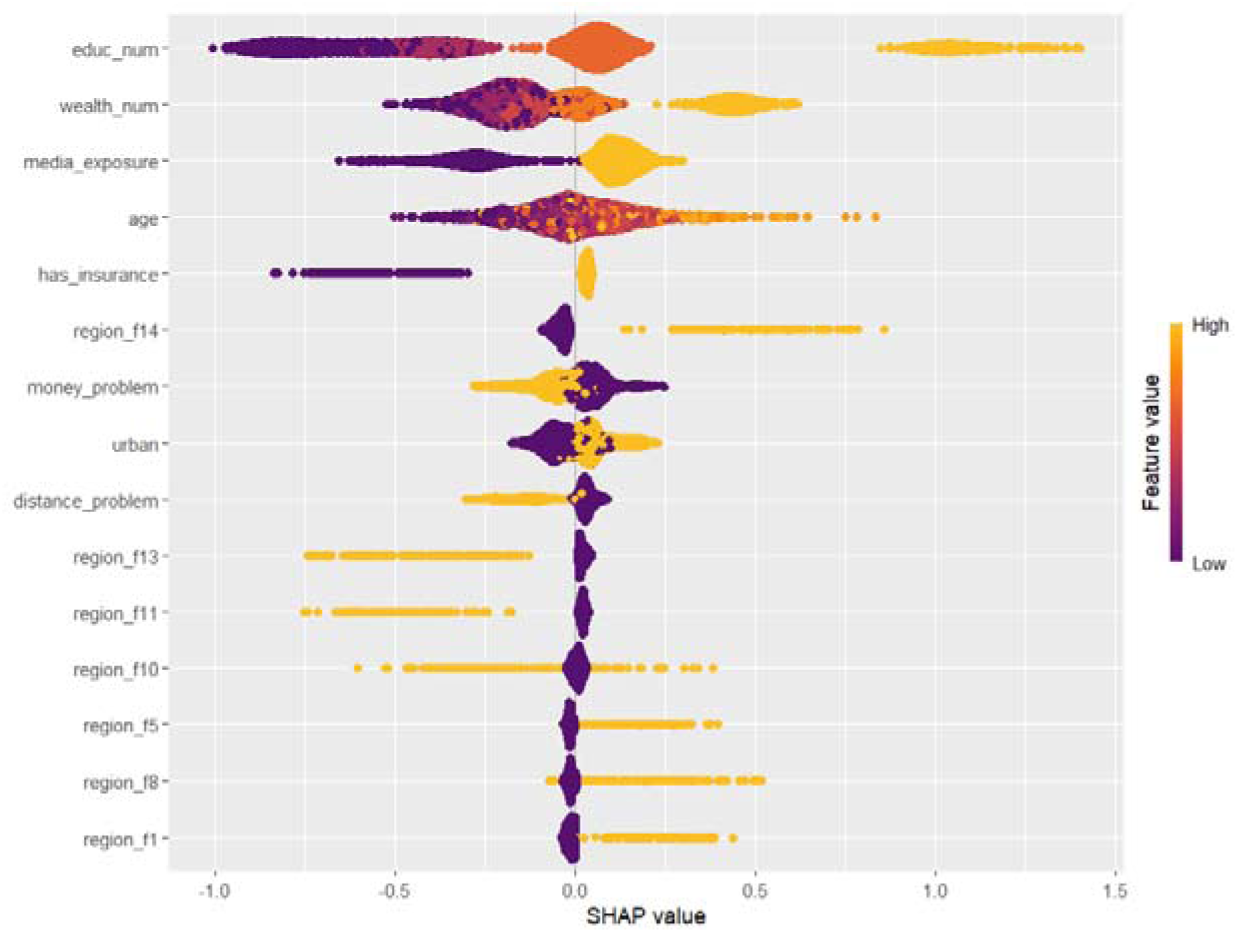
SHAP summary plot for the clinical breast examination model, random subsample of 2,000 women. Color denotes feature value (purple low, orange high); horizontal position denotes contribution to predicted screening probability.

### Concentration index and socioeconomic equity

Concentration curves for both outcomes lay below the diagonal of equality across almost the entire wealth distribution (Figure 8), indicating pro-rich concentration in both services. The ordinary concentration index was 0.256 for breast examination and 0.348 for cervical testing, which read alone would suggest cervical testing is the more unequal service. Because this index is bounded by 1 − μ for a binary outcome and the two outcomes have very different mean prevalence, the two figures are not on a common scale. The Erreygers correction gives 0.231 for breast examination against 0.095 for cervical testing (Table 3), reversing the comparison implied by the uncorrected index. Neither index carries a confidence interval in this analysis, so this reversal is reported as a point-estimate comparison rather than a statistically confirmed difference between the two outcomes; a bootstrap or analytic variance estimate for the difference in Erreygers indices would be needed to state this more firmly, and we recommend it for a revised analysis. A broadly similar sensitivity of concentration-index rankings to the choice of correction has been reported for cervical screening across several sub-Saharan African countries with differing baseline coverage [32].

**Figure 8.**
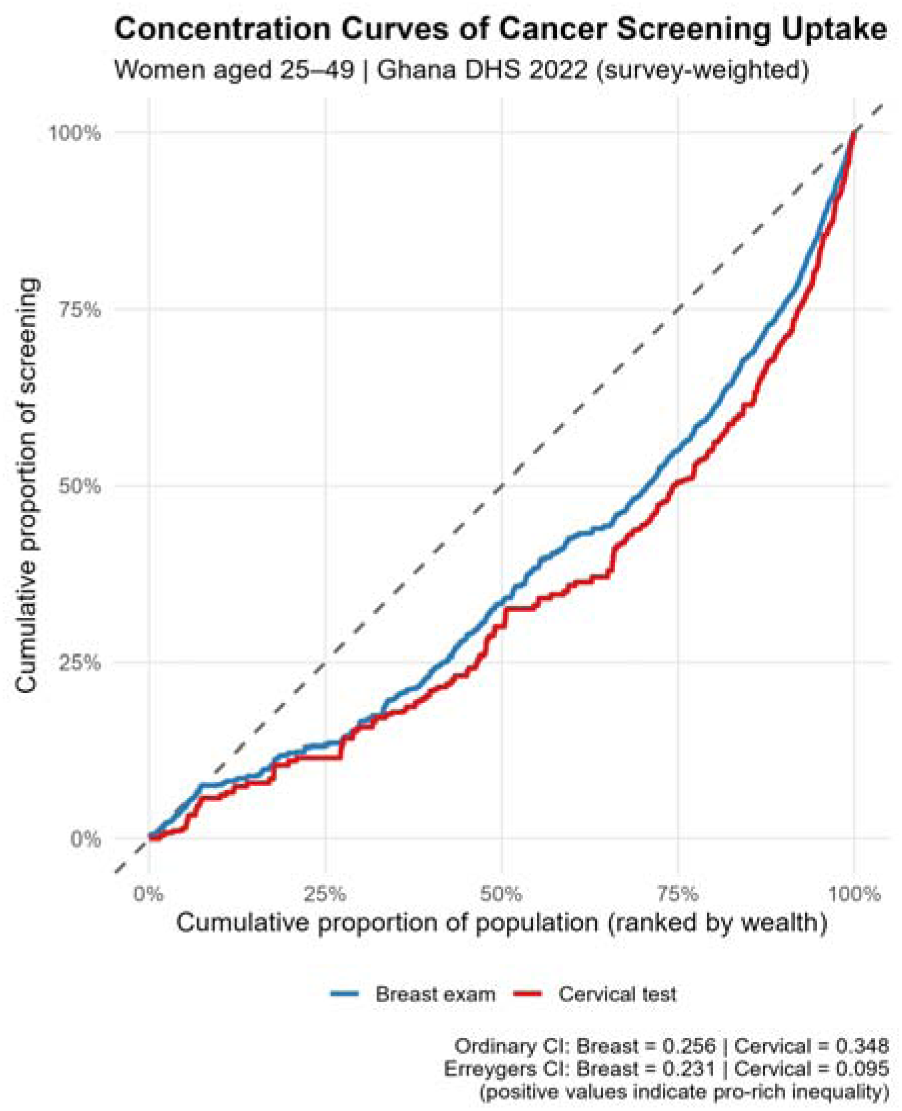
Concentration curves for clinical breast examination and cervical cancer testing, women 25-49 years, ranked by household wealth.

**Table 3.** Concentration indices for clinical breast examination and cervical cancer testing, women 25-49 years, ranked by household wealth. Positive values indicate pro-rich concentration; no confidence intervals are available (see Methods).

| Outcome | Mean prevalence | Ordinary concentration index | Erreygers concentration index |
| --- | --- | --- | --- |
| Clinical breast examination | 22.5% | 0.256 | 0.231 |
| Cervical cancer testing | 6.9% | 0.348 | 0.095 |

## Discussion

These results are consistent with, and extend, the pattern already reported for Ghana using the 2022 GDHS: education, wealth, urban residence, and to a smaller extent age and insurance status are associated with who receives clinical breast examination and cervical cancer testing, and both remain underused [1,2,7]. Our contribution is methodological rather than substantive, and we keep the two methodological strands separate in interpreting them. Cluster-level screening rates are significantly spatially autocorrelated for both outcomes, and Gi* analysis, read as an exploratory map rather than a confirmed set of hotspots given the lack of multiplicity correction, localizes that autocorrelation to specific areas rather than a smooth regional gradient. Separately, individual-level socioeconomic prediction of screening, validated with cluster-held-out folds to remove within-cluster leakage, reaches moderate discrimination for both outcomes; because the underlying models contain no geographic covariates, this predictive result speaks to how well socioeconomic composition predicts screening for women in unseen clusters, not to whether location itself adds predictive value beyond composition.

Treating both outcomes together, with matched diagnostics, surfaces two observations a single-outcome study would miss, both of which we state as associations rather than mechanisms. First, the spatial footprints of breast and cervical screening hotspots correlate but are not identical: cervical-testing hotspots reach further north than breast-examination hotspots, a pattern that fits the different delivery models the two services typically use (breast examination can be offered opportunistically during many kinds of clinical contact, while cervical testing more often needs a dedicated visit or campaign), a distinction also raised in facility-distance analyses of cervical screening across the wider region [9,33]; we have not, however, tested this delivery-model explanation directly. Second, the Erreygers-corrected concentration index reverses the ranking implied by the ordinary index: read on its own, cervical testing’s higher ordinary index (0.348 against 0.231) would suggest it is the more unequal service, but once both indices are placed on a common, prevalence-independent scale, breast examination appears more wealth-concentrated. We report this as a point-estimate reversal, since neither index carries a confidence interval in this analysis, and note that comparable sensitivity between the two indices has been documented for cervical screening across several sub-Saharan African countries with different baseline coverage [32], suggesting the pattern is not obviously an artifact specific to this dataset, though a formal statistical comparison would be needed to confirm it here.

For screening programs in Ghana, the hotspot maps offer a potentially more granular geography than regional tables, but we stop short of calling them a validated targeting tool. Apparent coldspot clusters for both outcomes fall mostly across the Northern, North East, Upper East, and Upper West regions, broadly consistent with the well-documented north-south gap in health infrastructure density, and Gi* also flags individual coldspot clusters inside otherwise better-performing regions and hotspot clusters inside lower-performing ones that a purely regional allocation rule would miss in both directions. The cluster-level risk ranking built from the fitted models is best described as a composition-based ranking of the sampled clusters’ predicted socioeconomic risk profile, not an independently validated spatial-prediction or targeting tool; it has not been compared against regional allocation on calibration, targeting accuracy, or decision utility, its uncertainty has not been quantified, and it has not been tested against clusters or areas outside the sampled data. We treat all three as prerequisites for any programmatic recommendation rather than as already demonstrated.

The feature-importance and SHAP results point to a related, practical consideration: because region indicators added comparatively little once individual-level education, wealth, and age were in the model, targeting based on regional averages alone will likely miscalibrate for individual clusters whose composition differs from their region’s average. Whether pairing regional allocation with cluster-level composition information, as explored for other health outcomes in Ghana using related methods [21,22], actually improves targeting performance in practice is an empirical question this study has not answered, and we recommend it as the natural next step rather than asserting the improvement here.

### Limitations

This analysis has several specific, addressable limitations. Because the GDHS is cross-sectional, all associations reported here, spatial and predictive alike, are descriptive rather than causal and cannot establish that living in a given cluster causes lower screening uptake rather than reflecting who lives there; we therefore describe spatial clustering as “associated with” or “compositionally patterned” by socioeconomic factors rather than “mediated” by them, and a formal mediation or spatial-decomposition analysis would be needed to support a stronger claim. Cluster-held-out cross-validation prevented women from the same cluster appearing in both training and test folds, but clusters were partitioned randomly rather than geographically, so the design tests generalization to unseen clusters within the sampled geography rather than extrapolation to distant or contiguous unsampled areas; a leave-region-out or spatially contiguous blocking scheme would give a stricter test. The XGBoost models used only individual socioeconomic and regional covariates rather than geographic coordinates and should be read as individual-level predictive models validated with cluster-aware resampling rather than as spatial machine-learning models that use location as a feature. GDHS cluster coordinates are also randomly displaced (up to 2 km in urban areas and 5–10 km in rural areas) to protect confidentiality, adding positional noise to the autocorrelation and hotspot analyses that we did not test with a displacement-sensitivity analysis. Case-wise deletion was used for the machine-learning sample without formally reporting the number of records excluded or how excluded women compared with those retained, which prevents an assessment of whether complete-case analysis introduced selection bias. Finally, hyperparameters were fixed a priori without nested tuning, early stopping and final performance evaluation shared the same held-out fold (introducing a small optimistic bias), calibration was not assessed, and no comparison against a survey-weighted logistic or multilevel regression baseline was performed, so this analysis does not establish that XGBoost adds predictive value over conventional regression for this problem.

### Conclusions

Breast and cervical cancer screening among Ghanaian women aged 25-49 years remains low and follows the education and wealth gradient already documented for this survey round. Beyond that, uptake of both services shows significant spatial autocorrelation at the level of individual survey clusters, with an exploratory set of hotspots and coldspots that regional reporting cannot show, and this clustering is associated with, though not formally demonstrated to be caused or mediated by, the socioeconomic composition of individual clusters. Individual-level machine-learning models validated with cluster-held-out resampling reach moderate discrimination using routine socioeconomic covariates, without geographic coordinates as inputs. A prevalence-corrected measure of concentration suggests, as a point estimate, that breast examination carries the larger wealth-related gap once the two outcomes’ different baseline rates are accounted for. Whether pairing Ghana’s cancer-screening equity monitoring with cluster-level spatial description improves on regional targeting in practice is a plausible but untested hypothesis raised by these results, and we present it as a direction for further, more rigorously validated work rather than as an established conclusion.

## List of abbreviations

AUC: Area under the (receiver operating characteristic) curve
DHS: Demographic and Health Surveys (Program)
GDHS: Ghana Demographic and Health Survey
GPS: Global Positioning System
ROC: Receiver operating characteristic
SHAP: Shapley additive explanations
STROBE: Strengthening the Reporting of Observational Studies in Epidemiology
WHO: World Health Organization
XGBoost: Extreme Gradient Boosting

## Declarations

### Ethics approval and consent to participate

This study is a secondary analysis of the 2022 Ghana Demographic and Health Survey. The original survey protocol, including its informed-consent procedures, was reviewed and approved by the Ghana Health Service Ethics Review Committee and the Institutional Review Board of ICF International (Rockville, Maryland, USA), the DHS Program’s implementing partner; written informed consent was obtained from all original participants by the DHS Program at the time of data collection. The present analysis used de-identified, publicly available secondary data obtained through registration with the DHS Program and involved no additional contact with participants; consistent with standard DHS secondary-analysis policy, it did not require separate institutional ethical review at the authors’ institutions.

### Consent for publication

Not applicable. This manuscript does not contain any individual person’s data, images, or other identifying information in any form.

### Availability of data and materials

The 2022 Ghana Demographic and Health Survey individual recode and linked, displaced geographic cluster coordinates analyzed in this study are available from the DHS Program [34] upon registration and approval of a standard data-use request, in accordance with the DHS Program’s data access and confidentiality policy; geographic data are subject to a separate geospatial data-use agreement. Derived cluster-level aggregates and analysis code for the survey-weighted, spatial, and machine-learning components of this study are available from the corresponding author on reasonable request; the authors intend to deposit a public code repository (data-cleaning, survey design, spatial, machine-learning, SHAP, and concentration-index scripts, with a documented package environment) ahead of formal submission.

### Authors’ contributions

A.H.S conceived the study, conducted the statistical and spatial analyses, and drafted the manuscript. A.I.S. contributed to the methodology, public health interpretation & discussion and reviewed the manuscript. O.A.I. contributed to the machine-learning analysis and reviewed the manuscript. All authors read and approved the final version.

## Data Availability

Availability of data and materials: The dataset used in this study is publicly available from the DHS Program upon registration and approved request at https://dhsprogram.com.

## Acknowledgements

The authors thank the DHS Program for providing access to the survey data used in this analysis.

## Authors’ information

Not applicable.

## References

1. Anaba EA, Alor SK, Badzi CD, Mbuwir CB, Muki B, Afaya A. Drivers of breast cancer and cervical cancer screening among women of reproductive age: insights from the Ghana Demographic and Health Survey. BMC Cancer. 2024;24(1). doi:10.1186/s12885-024-12697-6

2. Akonde M, Anyinka J, Abah GA, Gupta RD, Frempong B, Swenson CC. Factors associated with clinical breast examination and cervical cancer screening uptake in Ghanaian women, evidence from the 2022 Ghana Demographic and Health Survey. Afr Health Sci. 2025;25(3):123–135. doi:10.4314/ahs.v25i3.17

3. Wongnaah F, Aboagye RG, Storph RP, Seidu AA, Adnani QES, Ahinkorah BO. Breast cancer screening uptake and its associated factors among women in Ghana: evidence from the 2022 Demographic and Health Survey. Arch Public Health. 2025;83. doi:10.1186/s13690-025-01741-x

4. Sung H, Ferlay J, Siegel RL, Laversanne M, Soerjomataram I, Jemal A, et al. Global cancer statistics 2020: GLOBOCAN estimates of incidence and mortality worldwide for 36 cancers in 185 countries. CA Cancer J Clin. 2021;71(3):209–249. doi:10.3322/caac.21660

5. Bray F, Laversanne M, Sung H, Ferlay J, Siegel RL, Soerjomataram I, et al. Global cancer statistics 2022: GLOBOCAN estimates of incidence and mortality worldwide for 36 cancers in 185 countries. CA Cancer J Clin. 2024;74(3):229-263. doi:10.3322/caac.21834

6. World Health Organization. Global strategy to accelerate the elimination of cervical cancer as a public health problem. Geneva: World Health Organization; 2020.

7. Ayanore MA, Adjuik M, Ameko A, Kugbey N, Asampong R, Mensah D, et al. Self-reported breast and cervical cancer screening practices among women in Ghana: predictive factors and reproductive health policy implications from the WHO study on global AGEing and adult health. BMC Womens Health. 2020;20(1):1–10 doi:10.1186/s12905-020-01022-5

8. Gyembuzie FK, Wongnaah F, Aboagye RG, Storph RP, Seidu AA, Adnani QES, et al. Regional variations in breast cancer screening uptake and its associated factors in Ghana. Eur J Public Health. 2024;34(Suppl 3). doi:10.1093/eurpub/ckae144.1267

9. Dickson KS, Boateng ENK, Acquah E, Ayebeng C, Addo IY. Screening for cervical cancer among women in five countries in sub-Saharan Africa: analysis of the role played by distance to health facility and socio-demographic factors. BMC Health Serv Res. 2023;23. doi:10.1186/s12913-023-09055-w

10. Appiah RS, Boakye K, Appiah G, Aidoo AA, Acquah-Hagan G, Singh B, et al. Rural-urban variations in cervical cancer screening uptake among women in Ghana: evidence from the 2022 Ghana Demographic and Health Survey. BMC Womens Health. 2025;25. doi:10.1186/s12905-025-03802-3

11. Burgert CR, Colston J, Roy T, Zachary B. Geographic displacement procedure and georeferenced data release policy for the Demographic and Health Surveys. DHS Spatial Analysis Report No. 7. Rockville: ICF International; 2013.

12. Saaka S, Hambali MG. Factors associated with cervical cancer screening among women of reproductive age in Ghana. BMC Womens Health. 2024;24. doi:10.1186/s12905-024-03367-7

13. Kyei-Arthur F, Agyekum MW, Afrifa-Anane GF, Alhassan N, Kugbey N, Nyarko KM. Cervical cancer screening prevalence and predictors among women aged 25-49 years in Ghana: a cross-sectional study. Health Sci Rep. 2026;9. doi:10.1002/hsr2.71971

14. Chen T, Guestrin C. XGBoost: a scalable tree boosting system. In: Proceedings of the 22nd ACM SIGKDD International Conference on Knowledge Discovery and Data Mining; 2016. p. 785–794. doi:10.1145/2939672.2939785

15. Baykemagn ND, Aweke MN, Mesfin A, Baffa LD, Agimas MC, Abuhay H, et al. Identifying predictors of cervical cancer screening uptake in Sub-Saharan Africa using machine learning: cross-sectional study. JMIR Public Health Surveill. 2025;11. doi:10.2196/71677

16. Arage FG, Tadese ZB, Taye EA, Tsegaw TK, Abate TG, Alemu EA. Cervical cancer screening uptake and its associated factor in Sub-Saharan Africa: a machine learning approach. BMC Med Inform Decis Mak. 2025;25. doi:10.1186/s12911-025-03039-y

17. Kebede FB, Mare KU, Arage FG, Tiyou A, Teshale G, Kebede AZ, et al. Predicting lifetime cervical cancer screening among sexually active women in low-and middle-income countries using machine learning models: evidence from Demographic and Health Surveys. AJOG Glob Rep. 2026. doi:10.1016/j.xagr.2026.100669

18. Hashtarkhani S, Zhou Y, Kumsa F, White-Means S, Schwartz DL, Shaban-Nejad A. Analyzing geospatial and socioeconomic disparities in breast cancer screening among populations in the United States: machine learning approach. JMIR Cancer. 2025;11. doi:10.2196/59882

19. Barnils PN, Schüz B. Intersectional analysis of inequalities in self-reported breast cancer screening attendance using supervised machine learning and PROGRESS-Plus framework. Front Public Health. 2024;11. doi:10.3389/fpubh.2023.1332277

20. Roberts DR, Bahn V, Ciuti S, Boyce MS, Elith J, Guillera-Arroita G, et al. Cross-validation strategies for data with temporal, spatial, hierarchical, or phylogenetic structure. Ecography. 2017;40(8):913–929. doi:10.1111/ecog.02881

21. Ghanem VG. Spatial and machine learning analysis of district-level health insurance inequities in Ghana. Cureus. 2026;18. doi:10.7759/cureus.101984

22. Ghanem VG. Spatial machine learning and longitudinal analysis of skilled antenatal care access and fertility-related inequities in Ghana (1988-2022). 2026. doi:10.64898/2026.07.16.26358217

23. Bintabara D, Kamata CC, Mohamedi R, Basinda N. Spatial autocorrelation and determinants of low uptake of breast cancer screening among women of reproductive age: a mixed-effect multilevel analysis of Tanzanian population-based survey. PLoS One. 2025;20:e0338337. doi:10.1371/journal.pone.0338337

24. Salifu Y, Walana W, Lasong J, Yakubu M, Wobi EB, Torpey K. Young women’s healthcare screening behaviours and sexual autonomy in Ghana: a spatial distribution and socioeconomic inequality analysis of a large population-based survey. Front Reprod Health. 2026;8. doi:10.3389/frph.2026.1751165

25. Erreygers G. Correcting the concentration index. J Health Econ. 2009;28(2):504–515. doi:10.1016/j.jhealeco.2008.02.003

26. von Elm E, Altman DG, Egger M, Pocock SJ, Gøtzsche PC, Vandenbroucke JP. The Strengthening the Reporting of Observational Studies in Epidemiology (STROBE) statement: guidelines for reporting observational studies. Lancet. 2007;370(9596):1453–1457. doi:10.1016/S0140-6736(07)61602-X

27. Adzigbli LA, Aboagye RG, Adeleye K, Osborne A, Ahinkorah BO. Cervical cancer screening uptake and its predictors among women aged 30-49 in Ghana: providing evidence to support the World Health Organization’s cervical cancer elimination initiative. BMC Infect Dis. 2025;25. doi:10.1186/s12879-025-10485-6

28. Moran PAP. Notes on continuous stochastic phenomena. Biometrika. 1950;37(1/2):17–23. doi:10.2307/2332142

29. Getis A, Ord JK. The analysis of spatial association by use of distance statistics. Geogr Anal. 1992;24(3):189–206. doi:10.1111/j.1538-4632.1992.tb00261.x

30. Lundberg SM, Lee SI. A unified approach to interpreting model predictions. Adv Neural Inf Process Syst. 2017;30.

31. Wagstaff A, Paci P, van Doorslaer E. On the measurement of inequalities in health. Soc Sci Med. 1991;33(5):545–557. doi:10.1016/0277-9536(91)90212-U

32. Seifu BL, Negussie YM, Asnake AA, Asebe HA, Fente BM, Asmare Z, et al. Wealth-related inequalities of women’s cervical cancer screening in 11 Sub-Saharan African countries: evidence from a pooled decomposition analysis. Sci Rep. 2025;15. doi:10.1038/s41598-025-96347-2

33. Tsekpetse P, Salu S, Otoo DM, Dushime JF, Shiuma J, Oloo B, Ahinkorah BO. Rural-urban disparities in cervical cancer screening uptake and its predictors among women aged 30-49 years in Ghana: a multivariate decomposition analysis. BMC Womens Health. 2025;25. doi:10.1186/s12905-025-03962-2

34. DHS Program. The Demographic and Health Surveys (DHS) Program [Internet]. Rockville: ICF; [cited 2026 Aug 10]. Available from: https://dhsprogram.com

